# The citrate transporter SLC13A5 as a therapeutic target for kidney disease: evidence from Mendelian randomization to inform drug development

**DOI:** 10.1101/2023.08.29.23294760

**Authors:** Dipender Gill, Loukas Zagkos, Rubinder Gill, Thomas Benzing, Jens Jordan, Andreas L. Birkenfeld, Stephen Burgess, Grit Zahn

## Abstract

**Background:** Solute carrier family 13 member 5 (SLC13A5) is a Na -coupled citrate co-transporter that mediates the entry of extracellular citrate into the cytosol. SLC13A5 inhibition has been proposed as a therapeutic target for reducing progression of kidney disease via its effects on citrate metabolism. However, evidence of its efficacy in humans is limited. The aim of this study was to leverage the Mendelian randomization paradigm to gain insight into the effects of SLC13A5 inhibition in humans, towards prioritizing and informing clinical development efforts.

**Methods:** The primary Mendelian randomization analyses investigated the effect of SLC13A5 inhibition on measures of kidney function, including creatinine and cystatin C based measures of estimated glomerular filtration rate (creatinine-eGFR and cystatin C-eGFR), blood urea nitrogen (BUN), urine albumin-creatinine ratio (uACR), and risk of chronic kidney disease and microalbuminuria. Secondary analyses included a paired plasma and urine metabolome-wide association study, investigation of secondary traits related to SLC13A5 biology, a phenome-wide association study (PheWAS), and a proteome-wide association study. All analyses were compared to the effect of genetically predicted plasma citrate levels using variants selected from across the genome, and statistical sensitivity analyses robust to the inclusion of pleiotropic variants were also performed. Data were obtained from large scale genetic consortia and biobanks, with sample sizes ranging from 5,023 to 1,320,016 individuals.

**Results:** We found evidence of associations between genetically proxied SLC13A5 inhibition and higher creatinine-eGFR (p=0.002), cystatin C-eGFR (p=0.005), and lower BUN (p=3×10^-3^). Statistical sensitivity analyses robust to the inclusion of pleiotropic variants suggested that these effects may be a consequence of higher plasma citrate levels. There was no strong evidence of associations of genetically proxied SLC13A5 inhibition with uACR or risk of CKD or microalbuminuria. Secondary analyses also identified evidence of associations with higher plasma calcium levels (p=6×10^-13^) and lower fasting glucose (p=0.02). PheWAS did not identify any safety concerns.

**Conclusion:** This Mendelian randomization analysis provides human-centric insight to guide clinical development of an SLC13A5 inhibitor. We identify plasma calcium and citrate as biologically plausible biomarkers of target engagement, and plasma citrate as a potential biomarker of mechanism of action. Our human genetic evidence corroborates evidence from various animal models to support effects of SLC13A5 inhibition on improving kidney function. Further study in the form of early-stage clinical trials may now be warranted to help translate these findings towards improving patient care.

## Introduction

The kidney is a highly metabolically active organ. The provision of citric acid cycle intermediates has ameliorated renal tubular damage and progression of chronic renal injury in animal models [1–3]. Solute carrier family 13 member 5 (SLC13A5), which is primarily expressed in hepatocytes of the liver, is a membrane-bound Na -coupled co-transporter responsible for moving extracellular citrate into the cytosol [4,5]. Systemic SLC13A5 inhibition could augment citrate flux to the kidney, thereby supporting renal energy metabolism. Moreover, given the role of citrate in regulating glucose metabolism, lipid metabolism, and inflammation, SLC13A5 inhibition could indirectly affect renal health [6–13]. However, no SLC13A5 inhibitor has yet entered clinical study, and so evidence of its efficacy for ameliorating progression of kidney disease in humans is limited. Additionally, SLC13A5 shows species-specific effects [11,14–16].

Insights from human genetic data offer a powerful opportunity to prioritize and inform the design of clinical research. Given that learnings from such genetic analyses relate to the target organism, namely humans, they are well-placed to overcome some of the limitations of translating findings from animal models [17]. As genes code for proteins and proteins make up the majority of drug targets, it follows that naturally occurring variation in the genes coding for drug target proteins can be leveraged to inform on the effect of their pharmacological perturbation [18]. The random allocation of genetic variants at conception means that their association with clinical traits and phenotypes are less susceptible to the confounding factors and reverse causation bias that can hinder causal inference in traditional epidemiological study designs. The current availability of publicly accessible large-scale genetic association data also means that such a drug-target Mendelian randomization paradigm is a relatively fast and cost-effective approach for inferring the potential clinical effects of perturbing drug targets [19].

Given the pre-clinical evidence supporting potential therapeutic applications of SLC13A5 inhibition [10,12,13,20,21], we sought to investigate its effects using drug target Mendelian randomization. By utilizing the known effect of SLC13A5 inhibition on increasing plasma citrate levels to identify genetic instruments [10,22,23], our primary objective was to investigate its effects on parameters of kidney function. In secondary analyses, we aimed to unravel potential mechanisms of action, as well as evidence of broader effects on glucose and lipid metabolism, and inflammation. Such insights would thus serve to inform and prioritize clinical development efforts for SLC13A5 inhibitors.

## Methods

### Study design overview

Genetic instruments for SLC13A5 inhibition were first identified as minimally correlated (r <0.1) single-nucleotide polymorphisms (SNPs) within 200kB of the SLC13A5 gene (chromosome 17, base position 6,588,032 to 6,616,886 on reference panel GRCh37/hg19 by Ensembl) that associate with plasma citrate levels at a genome-wide significance level (p<5×10^-8^). Instrument validity was affirmed by performing Mendelian randomization to explore associations with plasma calcium levels, whose levels are postulated to be increased with SLC13A5 inhibition due to reduced sequestration in the bone [24]. Such an association would also support plasma calcium as a biomarker of target engagement for SLC13A5 inhibition.

Primary Mendelian randomization analyses were then performed to investigate the association of genetically proxied SLC13A5 inhibition with creatinine and cystatin C based measures of estimated glomerular filtration rate (eGFR), blood urea nitrogen (BUN), urine albumin-creatinine-ratio (uACR), and risk of microalbuminuria and chronic kidney disease (CKD).

To investigate whether any effect of SLC13A5 inhibition on parameters of kidney function may be attributable to metabolic effects in the plasma that consequently affect kidney function, metabolome-wide Mendelian randomization of the plasma and urine was undertaken for SLC13A5 inhibition. Evidence of concordant effects of SLC13A5 inhibition on biomarkers measured in the plasma and urine would be consistent with metabolic effects resulting in compensatory mechanisms in the kidney, whereas evidence of contrasting effects in the plasma and urine would suggest a process specific to the kidney underlying the effects.

Secondary Mendelian randomization analyses were performed to explore the human genetic evidence for effects of SLC13A5 inhibition on biomarkers of glucose and lipid metabolism, and inflammation, namely plasma low-density lipoprotein cholesterol (LDLc), high-density lipoprotein cholesterol (HDLc), triglycerides, fasting glucose, interleukin 6 (IL6) and C-reactive protein (CRP). Finally, given that SLC13A5 is predominantly expressed in the liver and is involved in glucose and lipid metabolism [10,12,20,25], we also investigated potential effects on liver fat.

In order to gain potential insight into mediating mechanisms and pathways implicated in the effects of SLC13A5 inhibition, we also performed a proteome-wide Mendelian randomization study. Finally, to investigate potential on-target adverse effects or novel indications, we performed a phenome-wide association study (PheWAS) [26].

To explore whether plasma citrate is a potential mediating mechanism for any associations identified, and thus could serve as a biomarker of mechanism of action, all analyses that considered genetically proxied SLC13A5 as the exposure were also repeated for genetically predicted plasma citrate levels, by selecting instruments from throughout the genome rather than confined to the SLC13A5 gene region.

A schematic figure depicting the overall study design is presented in Figure 1.

**Figure 1.**
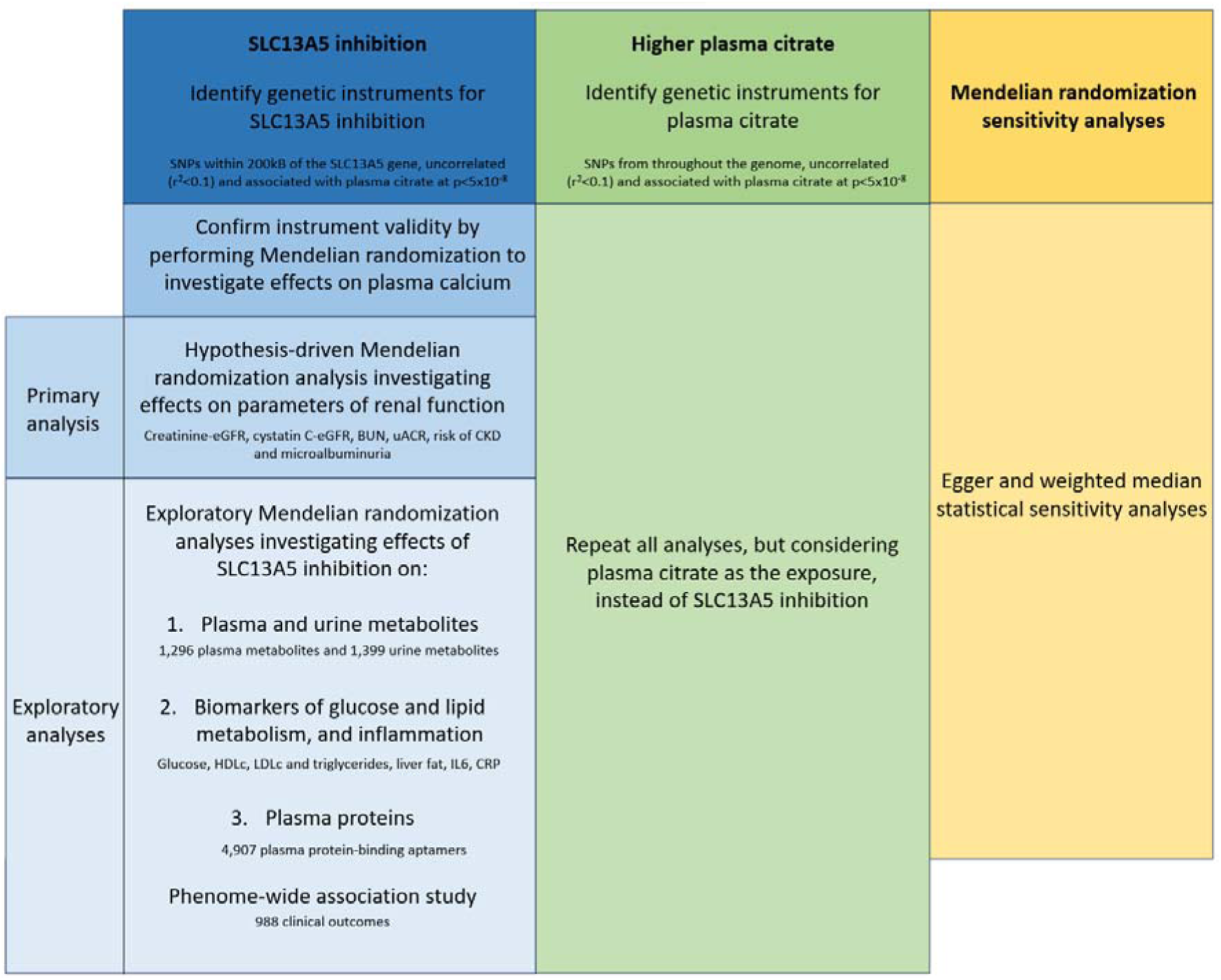
**Overview of the study design.** BUN: blood urea nitrogen, CRP:C-reactive protein, CKD: chronic kidney disease; eGFR: estimated glomerular filtration rate, HDLc: high-density lipoprotein cholesterol, IL6: interleukin 6; LDLc: low-density lipoprotein cholesterol, SNP: single-nucleotide polymorphism, uACR: urine albumin-creatinine ratio.

### Statistical analysis Instrument strength

Instrument strength was estimated by calculating the F-statistic for each SNP using the chi-squared approximation, which for each variant is the square of the SNP-exposure association estimate divided by the square of the SNP-exposure association estimate’s standard error [27].

### Mendelian randomization

The genome-wide association study summary data sources used for the Mendelian randomization analyses are presented in Table 1 [28–38]. Participant consent and ethical approval for all data were obtained in the original studies.

**Table 1.** Genome-wide association study summary data used for the Mendelian randomization analyses.

| Category | Trait | Unit | Population | Sample size | Citation |
| --- | --- | --- | --- | --- | --- |
| **Biomarker** | Plasma citrate | SD | European ancestry UK Biobank participants | 115,064 | [28] |
|  | Plasma calcium | SD | British ancestry UK Biobank participants | 361,194 | [29] |
| **Renal** | Creatinine-eGFR | SD of log | Multi-ancestry GWAS meta-analysis | 1,201,929 | [30] |
|  | Cystatin C-eGFR | SD of log |  |  |  |
|  | Blood urea nitrogen | SD of log |  |  |  |
|  | Urine albumin-creatinine ratio | SD of log | Multi-ancestry GWAS meta-analysis | 564,257 | [31] |
|  | Chronic kidney disease | Log OR | Multi-ancestry GWAS meta-analysis | 625,219 (64,164 cases) | [32] |
|  | Microalbuminuria | Log OR | Multi-ancestry GWAS meta-analysis | 348,954 (51,861 cases) | [31] |
| **Metabolome** | 1,296 plasma metabolites | SD | German Chronic Kidney Disease study | 5,023 | [33] |
|  | 1,399 urine metabolites | SD |  |  |  |
| **Lipid, glucose and inflammation** | Plasma low-density lipoprotein cholesterol | SD | European ancestry GWAS meta-analysis | 1,320,016 | [34] |
|  | Plasma high-density lipoprotein cholesterol | SD |  |  |  |
|  | Plasma triglycerides | SD |  |  |  |
|  | Fasting plasma glucose | mmol/l | European ancestry GWAS meta-analysis | 200,622 | [35] |
|  | Imaging-derived liver fat (MRI-PDFF) | SD | UK Biobank participants | 36,703 | [36] |
|  | Plasma interleukin 6 binding aptamer level | SD | deCODE Icelandic study | 35,559 | [37] |
|  | Plasma C-reactive protein level | SD of log | European ancestry GWAS | 427,367 | [38] |
| **Proteome** | 4,907 plasma protein-binding aptamers | SD | deCODE Icelandic study | 35,559 | [37] |

The full list of analysed metabolites and protein-binding aptamers are available in the Supplementary files. Full study details and access to GWAS summary data are provided in the original publications. BUN: blood urea nitrogen; eGFR: estimated glomerular filtration rate; GWAS: genome-wide association study; MRI-PDFF: magnetic resonance imaging derived proton density fat fraction; OR: odds ratio; SD: standard deviation; uACR: urine albumin-creatinine ratio.

The main Mendelian randomization analyses were performed using the random-effects inverse-variance weighted (IVW) method [39]. This meta-analyses the Wald ratio estimates for each instrument SNP using a random-effects inverse-variance model. After harmonising SNPs by their effect alleles, the Wald ratio estimate is calculated by dividing the SNP-exposure association estimate by the SNP-outcome association estimate. The standard error of the Wald ratio estimate was calculated using the propagation of error method.

Two Mendelian randomization statistical sensitivity analyses were employed, Egger and weighted median, which make distinct assumptions regarding the inclusion of pleiotropic variants. The Egger method regresses the SNP-outcome association estimates on the SNP-exposure association estimates [40]. The slope of the regression provides a Mendelian randomization estimate that is corrected for pleiotropic associations of the genetic variants and the intercept serves as a test for the presence of such pleiotropic associations, provided that the magnitudes of these pleiotropic associations are not correlated to instrument strength. The weighted median method orders the Wald ratio estimates for all instrument SNPs weighted by their precision, and selects the median value, with 95% confidence intervals (95% CIs) calculated by bootstrapping [41]. It provides an accurate Mendelian randomization estimate when more than half the information for the analysis comes from valid instruments. Heterogeneity in MR estimates derived from individual SNPs can also signify the presence of potential bias from pleiotropy and was assessed using the Cochran’s Q test [42]. All Mendelian randomization analyses were performed using the “MendelianRandomization” package for R statistical software [43].

### Phenome-wide association study

PheWAS was performed in the UK Biobank, a prospective cohort study initiated in 2006 that recruited more than 500,000 participants aged between 40 and 69 years [44]. Participants contributed phenotypic and genetic data, as well as biological samples, as previously reported [45]. UK Biobank has approval from the North West Multi-centre Research Ethics Committee and all participants provided appropriate consent.

PheWAS statistical analysis was performed by first constructing a weighted genetic risk score for the exposure using the same instrument SNPs as employed in the Mendelian randomization analysis. For each individual included in the analysis, the number of plasma citrate-increasing alleles for each instrument SNP were multiplied by the corresponding genetic association estimate with citrate for that SNP, before combining by addition. International Classification of Diseases versions 9 and 10 were used to ascertain cases in the UK Biobank Hospital Episode Statistics data, with diagnoses linked to the phenotype code (phecode) grouping system to facilitate classification of clinically relevant traits [46]. Logistic regression was performed for each phecode against the standardized genetic risk score for the exposure, adjusting for age, sex, and the first 10 genetic principal components of genetic ancestry. Only phecodes with 200 or more cases were included in the analysis, to avoid inclusion of outcomes with low statistical power. PheWAS estimates are reported per 1-SD higher standardized exposure genetic risk score. PheWAS was conducted using the “PheWAS” package of R statistical software [47].

### Statistical significance ascertainment

To account for testing of multiple outcomes in the primary Mendelian randomization analysis of renal outcomes, the Benjamini–Hochberg false discovery rate (FDR) 5% threshold was used. No statistical significance threshold was used for the other analyses, which were exploratory in nature. All presented p-values are uncorrected for multiple testing, unless otherwise stated.

### Comparison of SLC13A5 inhibition with higher plasma citrate levels by any mechanism

The Mendelian randomization and PheWAS analyses were also performed considering the exposure of plasma citrate levels, rather than SLC13A5 inhibition. Selection of genetic variants to serve as instruments and build the weighted genetic risk score for plasma citrate was undertaken using the same approach as for SLC13A5 inhibition, except that selection of variants was not confined to the SLC13A5 gene region.

### Correlation between plasma and urine metabolome-wide Mendelian randomization results

To measure the correlation between the plasma and urine metabolome-wide Mendelian randomization z-score results, comparison was restricted to all those metabolites analysed in both plasma and urine, and associated with genetically proxied SLC13A5 inhibition at FDR p < 0.2 in either analysis.

### Availability of code and data

R statistical software was used for all analyses. The statistical code used in this work is available upon reasonable request to the corresponding author. All genome-wide association study summary data are publicly available from the sources cited in Table 1. UK Biobank individual participant data are available upon appropriate application to the UK Biobank study. This work was reported using the “Strengthening the Reporting of Observational Studies in Epidemiology using Mendelian Randomization” (STROBE-MR) checklist (Supplementary checklist) [48].

## Results

A total of 13 SNPs were identified as instruments for SLC13A5 inhibition (Supplementary Table 1), all of which had F-statistics > 10, consistent with low risk of weak instrument bias that might affect the conclusions of Mendelian randomization analyses.

The main IVW Mendelian randomization identified positive associations of genetically proxied SLC13A5 inhibition with plasma calcium levels. Every 1-SD higher genetically proxied plasma citrate through SLC13A5 inhibition was associated with a 0.132 SD units higher plasma calcium level (95% CI 0.107 to 0.157, p=6×10^-13^).

Figure 2 depicts the associations of genetically proxied SLC13A5 inhibition with kidney function parameters from the main IVW Mendelian randomization analysis. There were statistically significant associations of genetically proxied SLC13A5 inhibition with higher creatinine and cystatin C based measures of eGFR (FDR adjusted p values = 0.006 and 0.01 respectively), and lower BUN (FDR adjusted p value = 0.002). There were no strong associations of genetically proxied SLC13A5 inhibition with uACR (FDR adjusted p value = 0.682), or risk of CKD (FDR adjusted p value = 0.516) or microalbuminuria (FDR adjusted p value = 0.682).

**Figure 2.**
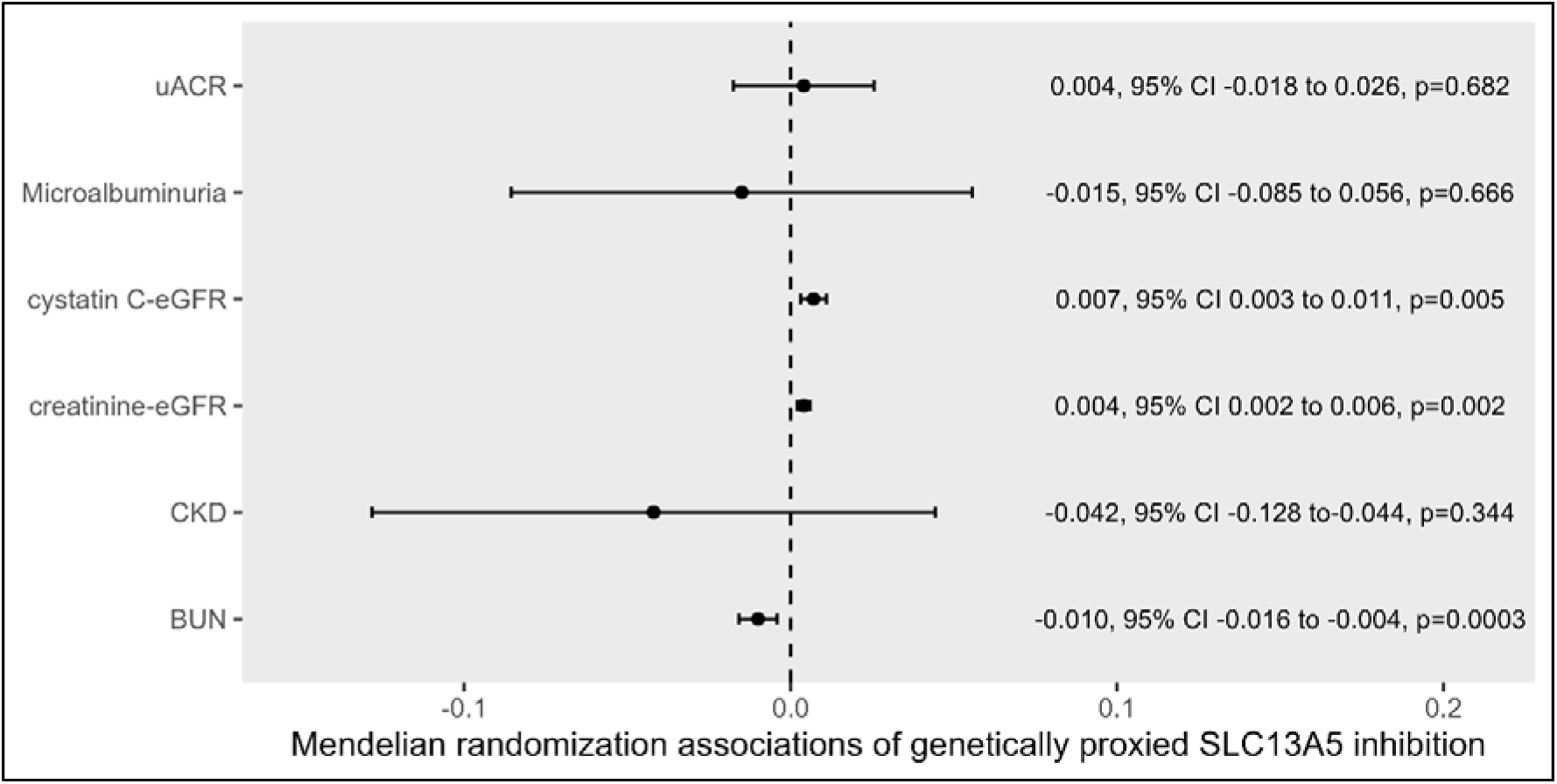
Mendelian randomization estimates for the association of genetically proxied SLC13A5 inhibition with kidney traits. Mendelian randomization estimates are scaled per 1-standard deviation (SD) increase in plasma citrate through genetically proxied SLC13A5 inhibition. The units for the BUN, creatinine-eGFR, cystatin C-eGFR and uACR are SD change in log transformed levels. The units for CKD and microalbuminuria are log odds ratio. Raw p values are presented. BUN: blood urea nitrogen, chronic kidney disease; eGFR: estimated glomerular filtration rate, urine albumin-creatinine ratio.

The results of the metabolome-wide Mendelian randomization analysis investigating the effects of SLC13A5 inhibition on plasma and urine metabolites are presented in Supplementary Tables 2 and 3. For the 7 metabolites present in both analyses and associated with genetically proxied SLC13A5 inhibition at FDR p < 0.2 in either analysis (citrate, aconitate, acetylspermidine, alpha-hydroxyisocaproate, propyl 4-hydroxybenzoate sulfate, (2-butoxyethoxy)acetic acid, 8- methoxykynurenate), the correlation coefficient of their z-scores in the analysis of plasma and urine was 0.78 (p=0.04).

The main IVW Mendelian randomization analysis results measuring the association of genetically proxied SLC13A5 inhibition with parameters of glucose and lipid metabolism, and inflammation are presented in Figure 3. In this hypothesis-generating analysis, there were no significant associations after adjusting for multiple testing, but some evidence to support possible associations of SLC13A5 inhibition with lower fasting glucose levels (-0.026 mmol/l per 1-SD higher plasma citrate through SCL13A5 inhibition, 95% CI -0.047 to 0.004, raw p=0.022). Neither proteome-wide Mendelian randomization (Supplementary Table 4) nor PheWAS (Supplementary Table 5) identified any significant associations for genetically proxied SLC13A5 inhibition after correcting for multiple testing.

**Figure 3.**
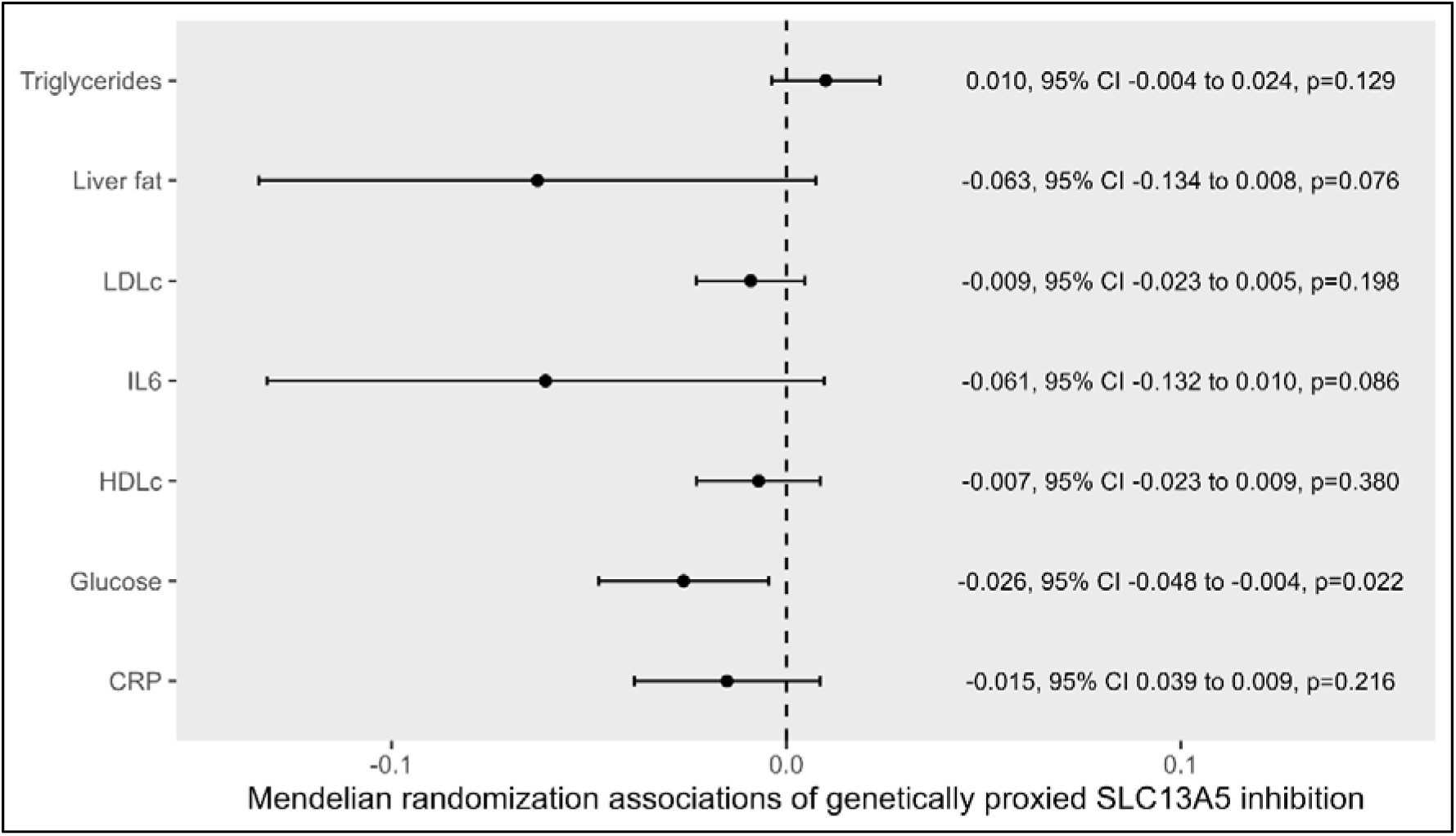
Mendelian randomization estimates for the association of genetically proxied SLC13A5 inhibition with biomarkers of glucose and lipid metabolism, and inflammation. Mendelian randomization estimates are scaled per 1-standard deviation (SD) increase in plasma citrate through genetically proxied SLC13A5 inhibition. The units for fasting glucose is mmol/l, the units for C-reactive protein (CRP) is standard deviation (SD) change of log transformed levels, and the remaining exposures are measured in SD units. HDLc: high-density lipoprotein cholesterol, IL6: interleukin 6, LDLc: low-density lipoprotein cholesterol.

All analyses were also performed considering higher plasma citrate through any mechanism as the exposure, instead of genetically proxied SLC13A5 inhibition (i.e., SNPs associated with higher plasma citrate selected as in the analysis for SLC13A5, but not restricted to 200kB within the SLC13A5 gene). The SNPs used as instruments for plasma citrate are presented in Supplementary Table 6, and the analysis results are presented in Supplementary Tables 7-10. Although the main IVW analysis did not identify evidence supporting associations of genetically predicted plasma citrate levels with eGFR or BUN, the Egger and weighted median sensitivity analyses that are more robust to the inclusion of pleiotropic SNPs were more supportive of such an association (Supplementary Table 11). This suggests that the genetic variants that affect plasma citrate are heterogeneous in the mechanisms by which they achieve this, and consistent with this, there was strong evidence of heterogeneity in all the IVW MR analyses undertaken with plasma citrate as the exposure (Supplementary Table 11).

## Discussion

Using large-scale genetic association data, we identified a genetic instrument for SLC13A5 inhibition that we leveraged in the Mendelian randomization paradigm to provide clinically translatable insights. Our main findings generated human genetic evidence that supports higher plasma citrate and calcium as biologically plausible biomarkers of target engagement, higher plasma citrate as a potential mechanism of action, and favourable effects on parameters of kidney function, namely creatinine and cystatin C based measures of eGFR, and BUN. Of note, we did not identify statistically significant evidence of effects of SLC13A5 inhibition on uACR nor risk of microalbuminuria. This discrepancy may be explained by these biomarkers measuring different domains of kidney function [49]. Of relevance, calcium citrate treatment in an acute kidney damage animal model resulted in significantly reduced levels of proteinuria [1]. The primary Mendelian randomization analysis also did not identify evidence of SLC13A5 inhibition having effects on CKD risk, which may in part be explained by the pathophysiological heterogeneity underlying CKD [50], and SLC13A5 inhibition potentially only being relevant to some of these mechanisms.

The correlation between the associations of genetically proxied SLC13A5 with the plasma and urine metabolomes is consistent with effects of SLC13A5 on the urine being downstream to effects on the plasma, and the kidney compensating plasma changes through altered renal excretion. This would also support that effects of SLC13A5 inhibition on kidney function may be occurring through metabolic effects originating outside the kidney. Secondary Mendelian randomization analyses supported potential effects of SLC13A5 inhibition on reducing fasting glucose. Given that SLC13A5 is predominantly expressed in the liver [25], these findings are consistent with direct hepatic effects. The lack of strong associations of genetically proxied SLC13A5 inhibition with clinical outcomes in PheWAS is reassuring regarding safety profile. This suggests smaller reductions in SLC13A5 activity are unlikely to recapitulate the autosomal-recessive epileptic encephalopathy phenotype observed with rare mutations affecting critical regions of the SLC13A5 gene [51].

SLC13A5 is a membrane-bound citrate transporter, and its inhibition results in reduced entry of plasma citrate into the cytosol [4,5]. This would be expected to have implications for cellular metabolism in cells expressing SLC13A5, with the higher circulating plasma citrate levels also potentially impacting metabolism systemically. Consistent with the results of our main genetic analyses, previous work has identified protective effects of citrate in various animal models of kidney disease [1,6,52,53]. Although the main IVW Mendelian randomization analysis investigating effects of plasma citrate levels did not identify evidence supporting effects on parameters of kidney function, there was strong evidence of heterogeneity in estimates generated by individual SNPs, consistent with the presence of pleiotropic effects that could bias the main IVW analyses [42]. Furthermore, the Egger and weighted median Mendelian randomization statistical sensitivity analyses, which can be more robust to the inclusion of pleiotropic variants, produced estimates more suggestive of favourable effects of higher plasma citrate on measures of kidney function. This observation may suggest that plasma citrate is a heterogeneous trait that is affected through several distinct pathways, and that at least some of these may favourably impact kidney function. This would be in-keeping with the favourable effects of SLC13A5 on kidney function occurring through its effects on plasma citrate. It has been reported that low citrate levels in urine are associated with various forms of kidney disease, and low urinary citrate is a biomarker for kidney disease progression [52,54–59]. Furthermore, preclinical studies with SLC13A5 knockout mice have supported effects on sympathoadrenal mechanisms [60], which may also contribute to nephroprotective effects of genetically proxied SLC13A5 inhibition.

There are several hypotheses for how citrate may exert its beneficial effects on kidney function. Generally, elevated plasma citrate levels are proposed to lead to increased excretion of citrate through the urine. Higher urinary citrate may in turn provide an additional energy supply for kidney cells, potentially enhancing their resilience and function. Furthermore, citrate may contribute to supporting the biosynthesis of essential molecules such as glucose, lipids, and amino acids. Additionally, it is known, due to its alkalinizing function citrate contributes to improved acid-base balance within the kidneys, promoting a favourable environment for cellular processes. Finally, urinary citrate’s solubilizing properties might offer protection against vascular calcification and the formation of kidney stones, thus safeguarding kidney health.

This work has several strengths. By leveraging large-scale genetic association data within the Mendelian randomization paradigm, we were able to rapidly perform several hypothesis-driven and hypothesis-free genetic analyses relevant to humans, and consequently generate insight related to drug development. Our findings represent numerous potential learnings, including highlighting higher plasma citrate and calcium as a biomarker of target engagement, higher plasma citrate as a biomarker of mechanism of action, efficacy for reducing progression of kidney disease, general safety profile, and potential effects on reducing fasting glucose. Further, the random allocation of genetic variants at conception means that these findings are less vulnerable to the confounding and reverse causation bias that can hinder causal inference in traditional epidemiological study designs [61].

This work should also be interpreted in the context of its limitations. The Mendelian randomization paradigm considers small lifelong effects of germline genetic variation. This is not the same as a discrete clinical intervention in later life, which may be of shorter duration but larger magnitude. Thus, it is possible that some of the findings of these genetic analyses may not predict what is observed with inhibition of SLC13A5 in clinical practice, particularly in quantitative terms. Further, some of our current analyses may also be limited by phenotypic definition and statistical power. While challenges in estimating the variance of drug target perturbation predicted by genetic variants means that it is not possible to perform conventional power calculations for drug target Mendelian randomization [62], some indication of the relative statistical power available for each analysis is apparent from the size of the 95% CIs. From this, it is clear that the analyses considering CKD had much less power than those considering measures of eGFR or BUN, for example. Of relevance, numerous preclinical mechanistic studies have demonstrated a role of SLC13A5 in favourably affecting hepatic lipid and glucose metabolism [10–13,20,21,63,64], and the limitations of our current analyses may explain the discrepancy in findings. The plasma and urine metabolome-wide Mendelian randomization analyses were undertaken using outcome data obtained from a CKD population and may thus be vulnerable to collider bias [65]. This is because stratifying on a collider can introduce associations with confounding factors. Our Mendelian randomization evidence supports effects of SLC13A5 inhibition on parameters of kidney function, in-keeping with CKD therefore representing a potential collider. While there are strategies available to help explore potential bias from this [66], they either require individual participant data, which were not available for the current Mendelian randomization analyses, or have other prohibitive limitations [67]. Finally, the genetic analyses undertaken in this work provide insight into the on-target effects of perturbing SLC13A5 [19], but cannot directly inform on the effects of any pharmacological agents used to inhibit SLC13A5, including their pharmacokinetic and pharmacodynamic properties, or off-target effects.

In summary, this Mendelian randomization analysis provides novel human-centric insight to guide the clinical development of an SLC13A5 inhibitor. We identify biologically plausible biomarkers of target engagement and mode of action, as well as genetic evidence supporting potential effects in improving kidney function, with higher plasma citrate levels a possible underlying mechanism. The null PheWAS findings are reassuring in that they do not identify any safety signals. Further study in the form of early-stage clinical trials may now be warranted to help translate these findings towards improving patient care.

## Supporting information

STROBE-MR checklist

## Data Availability

All genome-wide association study summary data are publicly available from the sources cited in Table 1. UK Biobank individual participant data are available upon appropriate application to the UK Biobank study.

## Acronyms

BUN: blood urea nitrogen
CI: confidence interval
CKD: chronic kidney disease
CRP: c-reactive protein
eGFR: estimated glomerular filtration rate
FDR: false-discovery rate
GWAS: genome-wide association study
HDLc: high-density lipoprotein cholesterol
IL6: interleukin-6
IVW: inverse-variance weighted
LDLc: low-density lipoprotein cholesterol
OR: odds ratio
PheWAS: phenome-wide association study
SD: standard deviation
SNP: single-nucleotide polymorphism
uACR: urine albumin-creatinine ratio

## Supplementary files

Supplementary checklist. Strengthening the Reporting of Observational Studies in Epidemiology using Mendelian Randomization checklist.

Supplementary Table 1. Single-nucleotide polymorphisms used as instruments for SLC13A5 inhibition. Genetic associations with plasma citrate are presented for variants within 200kB of the SLC13A5 gene. SNP: single-nucleotide polymorphism.

Supplementary Table 2. Metabolome-wide Mendelian randomization analysis investigating the associations of genetically proxied SLC13A5 inhibition with plasma metabolites. CI: confidence interval.

Supplementary Table 3. Metabolome-wide Mendelian randomization analysis investigating the associations of genetically proxied SLC13A5 inhibition with urine metabolites. CI: confidence interval.

Supplementary Table 4. Proteome-wide Mendelian randomization analysis investigating the associations of genetically proxied SLC13A5 inhibition with plasma protein-binding aptamers.

Supplementary Table 5. Phenome-wide association study investigating the associations of a standardised genetic risk score for SLC13A5 inhibition with clinical outcomes across the phenome.

Supplementary Table 6. Single-nucleotide polymorphisms used as instruments for plasma citrate levels, selected from throughout the genome. SNP: single-nucleotide polymorphism.

Supplementary Table 7. Metabolome-wide Mendelian randomization analysis investigating the associations of genetically predicted plasma citrate levels with plasma metabolites. CI: confidence interval.

Supplementary Table 8. Metabolome-wide Mendelian randomization analysis investigating the associations of genetically predicted plasma citrate levels with urine metabolites. CI: confidence interval.

Supplementary Table 9. Proteome-wide Mendelian randomization analysis investigating the associations of genetically predicted plasma citrate levels with plasma protein-binding aptamers.

Supplementary Table 10. Phenome-wide association study investigating the associations of a standardised genetic risk score for plasma citrate with clinical outcomes across the phenome.

Supplementary Table 11. Mendelian randomization statistical sensitivity analyses for biomarkers of renal function, glucose and lipid metabolism, and inflammation.

## References

[1] Gadola L, Noboa O, Marquez MN, Rodriguez MJ, Nin N, Boggia J, et al. Calcium citrate ameliorates the progression of chronic renal injury. Kidney Int 2004;65. 10.1111/j.1523-1755.2004.00496.x.

[2] Weinberg JM, Venkatachalam MA, Roeser NF, Saikumar P, Dong Z, Senter RA, et al. Anaerobic and aerobic pathways for salvage of proximal tubules from hypoxia-induced mitochondrial injury. Am J Physiol Renal Physiol 2000;279. 10.1152/ajprenal.2000.279.5.f927.

[3] Feldkamp T, Kribben A, Roeser NF, Senter RA, Weinberg JM. Accumulation of nonesterified fatty acids causes the sustained energetic deficit in kidney proximal tubules after hypoxia- reoxygenation. Am J Physiol Renal Physiol 2006;290. 10.1152/ajprenal.00305.2005.

[4] Zhang L, Hu W, Qiu Z, Li Z, Bian J. Opportunities and Challenges for Inhibitors Targeting Citrate Transport and Metabolism in Drug Discovery. J Med Chem 2023. 10.1021/acs.jmedchem.3c00179.

[5] Inoue K, Zhuang L, Maddox DM, Smith SB, Ganapathy V. Human sodium-coupled citrate transporter, the orthologue of Drosophila Indy, as a novel target for lithium action. Biochemical Journal 2003;374. 10.1042/BJ20030827.

[6] Bienholz A, Reis J, Sanli P, De Groot H, Petrat F, Guberina H, et al. Citrate shows protective effects on cardiovascular and renal function in ischemia-induced acute kidney injury. BMC Nephrol 2017;18. 10.1186/s12882-017-0546-1.

[7] Akhtar MJ, Khan SA, Kumar B, Chawla P, Bhatia R, Singh K. Role of sodium dependent SLC13 transporter inhibitors in various metabolic disorders. Mol Cell Biochem 2023;478. 10.1007/s11010-022-04618-7.

[8] Willmes DM, Kurzbach A, Henke C, Schumann T, Zahn G, Heifetz A, et al. The longevity gene INDY (I’m Not Dead Yet) in metabolic control: Potential as pharmacological target. Pharmacol Ther 2018;185. 10.1016/j.pharmthera.2017.10.003.

[9] Schumann T, König J, Henke C, Willmes DM, Bornstein SR, Jordan J, et al. Solute carrier transporters as potential targets for the treatment of metabolic disease. Pharmacol Rev 2020;72. 10.1124/pr.118.015735.

[10] Birkenfeld AL, Lee HY, Guebre-Egziabher F, Alves TC, Jurczak MJ, Jornayvaz FR, et al. Deletion of the mammalian INDY homolog mimics aspects of dietary restriction and protects against adiposity and insulin resistance in mice. Cell Metab 2011;14. 10.1016/j.cmet.2011.06.009.

[11] von Loeffelholz C, Lieske S, Neuschäfer-Rube F, Willmes DM, Raschzok N, Sauer IM, et al. The human longevity gene homolog INDY and interleukin-6 interact in hepatic lipid metabolism. Hepatology 2017;66. 10.1002/hep.29089.

[12] Zahn G, Willmes DM, El-Agroudy NN, Yarnold C, Jarjes-Pike R, Schaertl S, et al. A Novel and Cross-Species Active Mammalian INDY (NaCT) Inhibitor Ameliorates Hepatic Steatosis in Mice with Diet-Induced Obesity. Metabolites 2022;12. 10.3390/metabo12080732.

[13] Huard K, Brown J, Jones JC, Cabral S, Futatsugi K, Gorgoglione M, et al. Discovery and characterization of novel inhibitors of the sodium-coupled citrate transporter (NaCT or SLC13A5). Sci Rep 2015;5. 10.1038/srep17391.

[14] Kopel JJ, Bhutia YD, Sivaprakasam S, Ganapathy V. Consequences of NaCT/SLC13A5/mINDY deficiency: Good versus evil, separated only by the blood-brain barrier. Biochemical Journal 2021;478. 10.1042/BCJ20200877.

[15] Gopal E, Babu E, Ramachandran S, Bhutia YD, Prasad PD, Ganapathy V. Species-specific influence of lithium on the activity of SLC13A5 (NACT): Lithium-induced activation is specific for the transporter in primates. Journal of Pharmacology and Experimental Therapeutics 2015;353. 10.1124/jpet.114.221523.

[16] Inoue K, Zhuang L, Ganapathy V. Human Na+-coupled citrate transporter: Primary structure, genomic organization, and transport function. Biochem Biophys Res Commun 2002;299. 10.1016/S0006-291X(02)02669-4.

[17] Hingorani AD, Kuan V, Finan C, Kruger FA, Gaulton A, Chopade S, et al. Improving the odds of drug development success through human genomics: modelling study. Sci Rep 2019;9. 10.1038/s41598-019-54849-w.

[18] Gill D, Georgakis MK, Walker VM, Schmidt AF, Gkatzionis A, Freitag DF, et al. Mendelian randomization for studying the effects of perturbing drug targets. Wellcome Open Res 2021;6. 10.12688/wellcomeopenres.16544.2.

[19] Burgess S, Mason AM, Grant AJ, Slob EAW, Gkatzionis A, Zuber V, et al. Using genetic association data to guide drug discovery and development: Review of methods and applications. Am J Hum Genet 2023;110. 10.1016/j.ajhg.2022.12.017.

[20] Pesta DH, Perry RJ, Guebre-Egziabher F, Zhang D, Jurczak M, Fischer-Rosinsky A, et al. Prevention of diet-induced hepatic steatosis and hepatic insulin resistance by second generation antisense oligonucleotides targeted to the longevity gene mIndy (Slc13a5). Aging 2015;7. 10.18632/aging.100854.

[21] Brachs S, Winkel AF, Tang H, Birkenfeld AL, Brunner B, Jahn-Hofmann K, et al. Inhibition of citrate cotransporter Slc13a5/mINDY by RNAi improves hepatic insulin sensitivity and prevents diet-induced non-alcoholic fatty liver disease in mice. Mol Metab 2016;5. 10.1016/j.molmet.2016.08.004.

[22] Brown TL, Nye KL, Porter BE. Growth and overall health of patients with slc13a5 citrate transporter disorder. Metabolites 2021;11. 10.3390/metabo11110746.

[23] Bainbridge MN, Cooney E, Miller M, Kennedy AD, Wulff JE, Donti T, et al. Analyses of SLC13A5- epilepsy patients reveal perturbations of TCA cycle. Mol Genet Metab 2017;121. 10.1016/j.ymgme.2017.06.009.

[24] Dirckx N, Zhang Q, Chu EY, Tower RJ, Li Z, Guo S, et al. A specialized metabolic pathway partitions citrate in hydroxyapatite to impact mineralization of bones and teeth. Proc Natl Acad Sci U S A 2022;119. 10.1073/pnas.2212178119.

[25] Li Z, Wang H. Molecular mechanisms of the SLC13A5 gene transcription. Metabolites 2021;11. 10.3390/metabo11100706.

[26] Denny JC, Ritchie MD, Basford MA, Pulley JM, Bastarache L, Brown-Gentry K, et al. PheWAS: Demonstrating the feasibility of a phenome-wide scan to discover gene-disease associations. Bioinformatics 2010;26. 10.1093/bioinformatics/btq126.

[27] Li B, Martin EB. An approximation to the F distribution using the chi-square distribution. Comput Stat Data Anal 2002;40. 10.1016/S0167-9473(01)00097-4.

28. [28] Elsworth B, Lyon M, Alexander T, Liu Y, Matthews P, Hallett J, et al. The MRC IEU OpenGWAS data infrastructure. BioRxiv 2020.

29. [29] Neale Lab. GWAS of UK Biobank biomarker measurements 2023.

[30] Stanzick KJ, Li Y, Schlosser P, Gorski M, Wuttke M, Thomas LF, et al. Discovery and prioritization of variants and genes for kidney function in >1.2 million individuals. Nat Commun 2021;12. 10.1038/s41467-021-24491-0.

[31] Teumer A, Li Y, Ghasemi S, Prins BP, Wuttke M, Hermle T, et al. Genome-wide association meta-analyses and fine-mapping elucidate pathways influencing albuminuria. Nat Commun 2019;10. 10.1038/s41467-019-11576-0.

[32] Wuttke M, Li Y, Li M, Sieber KB, Feitosa MF, Gorski M, et al. A catalog of genetic loci associated with kidney function from analyses of a million individuals. Nat Genet 2019;51. 10.1038/s41588-019-0407-x.

[33] Schlosser P, Scherer N, Grundner-Culemann F, Monteiro-Martins S, Haug S, Steinbrenner I, et al. Genetic studies of paired metabolomes reveal enzymatic and transport processes at the interface of plasma and urine. Nat Genet 2023;55. 10.1038/s41588-023-01409-8.

[34] Graham SE, Clarke SL, Wu KHH, Kanoni S, Zajac GJM, Ramdas S, et al. The power of genetic diversity in genome-wide association studies of lipids. Nature 2021;600. 10.1038/s41586-021-04064-3.

[35] Chen J, Spracklen CN, Marenne G, Varshney A, Corbin LJ, Luan J, et al. The trans-ancestral genomic architecture of glycemic traits. Nat Genet 2021;53. 10.1038/s41588-021-00852-9.

[36] Haas ME, Pirruccello JP, Friedman SN, Wang M, Emdin CA, Ajmera VH, et al. Machine learning enables new insights into genetic contributions to liver fat accumulation. Cell Genomics 2021;1. 10.1016/j.xgen.2021.100066.

[37] Ferkingstad E, Sulem P, Atlason BA, Sveinbjornsson G, Magnusson MI, Styrmisdottir EL, et al. Large-scale integration of the plasma proteome with genetics and disease. Nat Genet 2021;53. 10.1038/s41588-021-00978-w.

[38] Said S, Pazoki R, Karhunen V, Võsa U, Ligthart S, Bodinier B, et al. Genetic analysis of over half a million people characterises C-reactive protein loci. Nat Commun 2022;13. 10.1038/s41467-022-29650-5.

[39] Burgess S, Butterworth A, Thompson SG. Mendelian randomization analysis with multiple genetic variants using summarized data. Genet Epidemiol 2013;37:658–65. 10.1002/gepi.21758.

[40] Bowden J, Davey Smith G, Burgess S. Mendelian randomization with invalid instruments: effect estimation and bias detection through Egger regression. Int J Epidemiol 2015;44:512– 25. 10.1093/ije/dyv080.

[41] Bowden J, Davey Smith G, Haycock PC, Burgess S. Consistent Estimation in Mendelian Randomization with Some Invalid Instruments Using a Weighted Median Estimator. Genet Epidemiol 2016;40:304–14. 10.1002/gepi.21965.

[42] Greco M F Del, Minelli C, Sheehan NA, Thompson JR. Detecting pleiotropy in Mendelian randomisation studies with summary data and a continuous outcome. Stat Med 2015;34. 10.1002/sim.6522.

[43] Yavorska OO, Burgess S. MendelianRandomization: An R package for performing Mendelian randomization analyses using summarized data. Int J Epidemiol 2017;46. 10.1093/ije/dyx034.

[44] Sudlow C, Gallacher J, Allen N, Beral V, Burton P, Danesh J, et al. UK Biobank: An Open Access Resource for Identifying the Causes of a Wide Range of Complex Diseases of Middle and Old Age. PLoS Med 2015;12. 10.1371/journal.pmed.1001779.

[45] Bycroft C, Freeman C, Petkova D, Band G, Elliott LT, Sharp K, et al. The UK Biobank resource with deep phenotyping and genomic data. Nature 2018;562. 10.1038/s41586-018-0579-z.

[46] Wu P, Gifford A, Meng X, Li X, Campbell H, Varley T, et al. Mapping ICD-10 and ICD-10-CM Codes to phecodes: Workflow development and initial evaluation. JMIR Med Inform 2019;7. 10.2196/14325.

[47] Carroll RJ, Bastarache L, Denny JC. R PheWAS: Data analysis and plotting tools for phenome- wide association studies in the R environment. Bioinformatics 2014;30. 10.1093/bioinformatics/btu197.

[48] Skrivankova VW, Richmond RC, Woolf BAR, Yarmolinsky J, Davies NM, Swanson SA, et al. Strengthening the Reporting of Observational Studies in Epidemiology Using Mendelian Randomization: The STROBE-MR Statement. JAMA - Journal of the American Medical Association 2021;326. 10.1001/jama.2021.18236.

[49] Lopez-Giacoman S. Biomarkers in chronic kidney disease, from kidney function to kidney damage. World J Nephrol 2015;4. 10.5527/wjn.v4.i1.57.

[50] Matovinović MS. 1. Pathophysiology and Classification of Kidney Diseases. EJIFCC 2009;20.

[51] Thevenon J, Milh M, Feillet F, St-Onge J, Duffourd Y, Jugé C, et al. Mutations in SLC13A5 cause autosomal-recessive epileptic encephalopathy with seizure onset in the first days of Life. Am J Hum Genet 2014;95. 10.1016/j.ajhg.2014.06.006.

[52] Rocha DR, Xue L, Gomes Sousa HM, Carvalho Matos AC, Hoorn EJ, Salih M, et al. Urinary Citrate Is Associated with Kidney Outcomes in Early Polycystic Kidney Disease. Kidney360 2022;3. 10.34067/KID.0004772022.

[53] Tanner GA, Tanner JA. Citrate therapy for polycystic kidney disease in rats. Kidney Int 2000;58. 10.1111/j.1523-1755.2000.00357.x.

[54] Posada-Ayala M, Zubiri I, Martin-Lorenzo M, Sanz-Maroto A, Molero D, Gonzalez-Calero L, et al. Identification of a urine metabolomic signature in patients with advanced-stage chronic kidney disease. Kidney Int 2014;85. 10.1038/ki.2013.328.

[55] Hallan S, Afkarian M, Zelnick LR, Kestenbaum B, Sharma S, Saito R, et al. Metabolomics and Gene Expression Analysis Reveal Down-regulation of the Citric Acid (TCA) Cycle in Non- diabetic CKD Patients. EBioMedicine 2017;26. 10.1016/j.ebiom.2017.10.027.

[56] Liu JJ, Liu S, Gurung RL, Ching J, Kovalik JP, Tan TY, et al. Urine tricarboxylic acid cycle metabolites predict progressive chronic kidney disease in type 2 diabetes. Journal of Clinical Endocrinology and Metabolism 2018;103. 10.1210/jc.2018-00947.

[57] Goraya N, Simoni J, Sager LN, Mamun A, Madias NE, Wesson DE. Urine citrate excretion identifies changes in acid retention as eGFR declines in patients with chronic kidney disease. Am J Physiol Renal Physiol 2019;317. 10.1152/ajprenal.00044.2019.

[58] Mutter S, Valo E, Aittomäki V, Nybo K, Raivonen L, Thorn LM, et al. Urinary metabolite profiling and risk of progression of diabetic nephropathy in 2670 individuals with type 1 diabetes. Diabetologia 2022;65. 10.1007/s00125-021-05584-3.

[59] Domrongkitchaiporn S, Stitchantrakul W, Kochakarn W. Causes of Hypocitraturia in Recurrent Calcium Stone Formers: Focusing on Urinary Potassium Excretion. American Journal of Kidney Diseases 2006;48. 10.1053/j.ajkd.2006.06.008.

[60] Willmes DM, Daniels M, Kurzbach A, Lieske S, Bechmann N, Schumann T, et al. The longevity gene mIndy (I’m Not Dead, Yet) affects blood pressure through sympathoadrenal mechanisms. JCI Insight 2021;6. 10.1172/jci.insight.136083.

[61] Burgess S, Butterworth A, Malarstig A, Thompson SG. Use of Mendelian randomisation to assess potential benefit of clinical intervention. BMJ 2012;345. 10.1136/bmj.e7325.

[62] Burgess S. Sample size and power calculations in Mendelian randomization with a single instrumental variable and a binary outcome. Int J Epidemiol 2014;43. 10.1093/ije/dyu005.

[63] Neuschäfer-Rube F, Schraplau A, Schewe B, Lieske S, Krützfeldt JM, Ringel S, et al. Arylhydrocarbon receptor-dependent mIndy (Slc13a5) induction as possible contributor to benzo[a]pyrene-induced lipid accumulation in hepatocytes. Toxicology 2015;337. 10.1016/j.tox.2015.08.007.

[64] Li L, Li H, Garzel B, Yang H, Sueyoshi T, Li Q, et al. SLC13A5 Is a novel transcriptional target of the pregnane x receptor and sensitizes drug-induced steatosis in human liver. Mol Pharmacol 2015;87. 10.1124/mol.114.097287.

[65] Paternoster L, Tilling K, Davey Smith G. Genetic epidemiology and Mendelian randomization for informing disease therapeutics: Conceptual and methodological challenges. PLoS Genet 2017;13. 10.1371/journal.pgen.1006944.

[66] Mitchell RE, Hartley AE, Walker VM, Gkatzionis A, Yarmolinsky J, Bell JA, et al. Strategies to investigate and mitigate collider bias in genetic and Mendelian randomisation studies of disease progression. PLoS Genet 2023;19. 10.1371/journal.pgen.1010596.

[67] Cai S, Allen RJ, Wain L V, Dudbridge F. Reassessing the association of MUC5B with survival in idiopathic pulmonary fibrosis. Ann Hum Genet 2023;87:248–53. 10.1111/ahg.12522.

